# The Influence of Phenotyping Method on Structural Neuroimaging Associations with Depression in UK Biobank

**DOI:** 10.1101/2020.12.18.20248488

**Authors:** Mathew A. Harris, Simon R. Cox, Laura de Nooij, Miruna C. Barbu, Mark J. Adams, Xueyi Shen, Ian J. Deary, Stephen M. Lawrie, Andrew M. McIntosh, Heather C. Whalley

**Affiliations:** Division of Psychiatry, University of Edinburgh, Edinburgh, UK; Department of Psychology, University of Edinburgh, Edinburgh, UK

**Keywords:** Depression, structural neuroimaging, levels of phenotyping, grey matter, white matter integrity, UK Biobank

## Abstract

**Background:** Depression is assessed in many different ways, with large population studies often relying on minimal phenotyping approaches. Genetic results suggest that more formal clinical diagnoses and simpler self-report measures of depression show some core similarities, but also important differences. It is not yet clear whether this is also the case for neuroimaging measures.

**Methods:** We studied 39,300 UK Biobank imaging participants (20,701 female; aged 44.6 to 82.3 years, M = 64.1, SD = 7.5) with structural neuroimaging (T1 and DTI) and depression data. Depression phenotypes included a minmal single-item self-report measure, an intermediate symptom-based measure of ‘probable’ depression, and a more clinically robust measure based on DSM-IV criteria. We tested i) associations between brain structural measures and each depression phenotype, and ii) the effects of depression phenotype on these associations.

**Results:** Small depression-brain structure associations (β < 0.1) were significant after FDR correction for many global and regional metrics for all three phenotypes. The most consistent imaging associations across depression phenotypes were for measures of white matter integrity. There were small but significant effects of phenotype definition primarily for cortical thickness, which showed stronger negative associations with Self-reported Depression than the symptom-based measures.

**Conclusion:** Similar to previous genetic studies, we found some consistent associations indicating a core component of depression across phenotypes, and some additional associations that were phenotype-specific. Although these specific results did not relate to depth of phenotyping as expected, effects of phenotype definition are still an important consideration for future depression research.

## Introduction

Major depressive disorder (MDD) affects a substantial and increasing proportion of the population (Kessler et al., 2007; Hidaka, 2012). There are currently no objective biomarkers to guide diagnosis, which instead relies on the time-consuming subjective assessment of a range of possible symptoms (Gallo & Rabins, 1999). Several diagnostic tools are available, along with a variety of assessments adapted for research purposes. The range of available measures all differ in how they define depression, yet many of these definitions are used interchangeably. This may underlie inconsistencies across studies, as studying different definitions of depression could mean focusing on quite different aspects of the disorder. In the context of the current move towards larger community-based studies, detailed face-to-face clinical assessments are often impractical, with simpler self-report and questionnaire-based measures being favoured instead. The implications for the field of depression research are not yet clear, and better understanding of the impact on results of different definitions and levels of phenotyping is imperative.

Howard et al. (2018) recently studied genetic contributions to depression in a very large sample of over 320,000 UK Biobank participants. Importantly, the study assessed genome-wide associations for multiple definitions of lifetime depression based on: i) a broad classification derived from minimal phenotyping of self-reported treatment seeking for ‘nerves, anxiety or depression’, ii) an intermediate phenotype of ‘probable’ depression derived from responses to several self-report mental health questionnaire items, and iii) a clinical MDD phenotype derived from diagnosis by doctors while in hospital. Interestingly, all phenotype definitions showed similar genetic associations to one another and to previous results, suggesting that each related to an overlapping core component of depression. However, there were also a number of significant genetic associations specific to each phenotype, suggesting that different definitions might reflect different underlying constructs, with different aetiologies. In another study of the same sample, Cai et al. (2018) found that depression heritability estimates and genetic associations also varied according to level of phenotyping, with single-item self-report measures of depression more closely related to neuroticism than to clinically defined MDD. We hypothesised that similar differences in relation to depth of phenotyping would be observed for associations with structural neuroimaging measures.

A considerable amount of research has focused on associations between depression and neuroimaging-derived measures of the brain. Depression-related structural differences have been observed in a range of areas (Drevets et al., 2008; Koolschijn et al., 2009; Grieve et al., 2013), but the greatest effects are reductions in the hippocampus (Videbech & Ravnkilde, 2004; Schmaal et al., 2016) and regions of prefrontal cortex (Lorenzetti et al., 2009; Belleau et al., 2019). Some studies have shown increases in certain regions, such as amygdala (Frodl et al., 2002; Lange & Irle, 2004) and anterior cingulate cortex (Ancelin et al., 2019), although these may represent confounding effects of medication rather than depression itself (Hamilton et al., 2008). The use of different depression definitions may also account for some of the variability in results across studies. Measures of white matter integrity also show associations with depression (McIntosh et al., 2013; Shen et al., 2017), indicating reduced connectivity, particularly between frontal and limbic regions (Korgaonkar et al., 2011; Ouyang et al., 2011). Functional neuroimaging findings provide further support for reduced connectivity in depression (Wang et al., 2012; Mulders et al., 2015; Brakowski et al., 2017), which would be consistent with lower white matter integrity. It is essential to address the potential impact of variable and often coarse assessment of MDD, especially given the phenotypic and genetic evidence that different definitions may reflect partially separable constructs.

We therefore sought to explicitly test the effects of depression phenotyping on associations with structural neuroimaging features. The present study used definitions at three phenotyping levels, and both grey and white matter structural metrics for UK Biobank imaging participants. We tested i) depression associations with measures of cortical regions, subcortical structures and white matter tracts for each of the three depression phenotypes, and ii) the effects of phenotype definition on these associations. We hypothesised i) reductions in grey matter size and white matter integrity among depressed subjects in a range of areas described above, and ii) that associations between depression and structural brain metrics would vary considerably across levels of phenotyping, with greater effect sizes for more in-depth phenotypes. These results would indicate whether associations between depression and neuroimaging measures depend on how depression is phenotyped or defined, relevant for future large-scale studies of the disorder.

## Methods

### Participants

UK Biobank (http://www/ukbiobank.ac.uk) includes data on 503,325 members of the general UK population, recruited between 2006 and 2010 (Allen et al., 2012). Participants originally provided information on a wide range of health, lifestyle, environment and other variables, and have provided further data at subsequent follow-ups. The present study focused on the first 39,300 of these participants for whom structural neuroimaging data were made available. Participants with neurological disorders (including schizophrenia, dementia, Parkinson’s disease and multiple sclerosis) were excluded from all analyses. The number of remaining subjects who also provided data on lifetime depression ranged from 15,079 to 26,875, depending on the depression phenotype. Descriptive statistics are reported in *Table 1*.

**Table 1.** Descriptive statistics for each depression phenotype.

|  | Self-reported<br>Depression | Probable<br>Depression | CIDI-assessed<br>MDD |
| --- | --- | --- | --- |
| Subjects (N, %) | 26,875 (68.4) | 15,079 (38.4) | 19,430 (49.4) |
| Age (years; M, SD) | 64.3 (7.6) | 63.3 (7.5) | 64.2 (7.5) |
| *Sex* |  |  |  |
| Males (N, %) | 13,977 (52.0) | 7,230 (47.9) | 9,165 (47.2) |
| Females (N, %) | 12,898 (48.0) | 7,849 (52.1) | 10,265 (52.8) |
| *Depression* |  |  |  |
| Cases (N, %) | 2,561 (9.5) | 6,787 (45.0) | 6,456 (33.2) |
| Controls (N, %) | 24,314 (90.5) | 8,292 (55.0) | 12,974 (66.8) |
| Neuroticism score (M, SD) | 3.3 (3.0) | 3.8 (3.2) | 3.4 (3.1) |
| Global CT (mm; M, SD) | 2.67 (0.11) | 2.67 (0.11) | 2.67 (0.11) |
| Total CSA (mm<sup>2</sup>; M, SD) | 169,880 (15,409) | 169,498 (15,308) | 169,399 (15,276) |
| Total CV (mm<sup>3</sup>; M, SD) | 500,279 (47,867) | 499,502 (47,575) | 498,866 (47,143) |
| Total SCV (mm<sup>3</sup>; M, SD) | 188,547 (169,19) | 188,073 (16,894) | 187,912 (16,691) |
| Global FA (M, SD) | 0.011 (1.002) | 0.030 (0.990) | 0.016 (0.992) |
| Global MD (M, SD) | 0.018 (1.003) | -0.002 (0.988) | -0.001 (0.992) |

*Notes.* CIDI = Composite International Diagnostic Interview (short form); MDD = Major Depressive Disorder; CT = cortical thickness; CSA = cortical surface area; CV = cortical volume; SCV = subcortical volume; FA = fractional anisotropy; MD = mean diffusivity. Descriptive statistics for secondary phenotypes are reported in supplementary materials (*Table S1*).

### Depression phenotypes

As the aim of this study was to assess the effect of different definitions of depression on underlying imaging features, depression was phenotypically defined in multiple ways. We focused primarily on three of the five definitions assessed previously by Cai et al. (2018): a minimal phenotype, Self-reported Depression, an intermediate phenotype, Probable Depression, and a more clinical definition, CIDI-assessed MDD. These phenotypes are described in further detail below, and numbers of cases and controls included by each definition are reported in *Table 1*. The same details for secondary phenotypes also included in supplementary materials (*Table S1*).

#### Minimal phenotypes

Our primary minimal phenotype, Self-reported depression, was based on whether participants did (cases) or did not (controls) specifically report a past diagnosis of depression when asked to provide details of any physical or mental disorders they had ever been diagnosed with. This assessment took place at the same time as neuroimaging. A second simple measure, Self-reported Treatment, is also reported in supplementary materials.

#### Intermediate phenotypes

An approximate measure of depression was previously created for UK Biobank participants (Smith et al., 2013), in lieu of more reliable clinical assessments. This was derived from a subset of questions on mental health, administered at each UK Biobank assessment by touchscreen. Questions relevant to depression included those on incidence and duration of previous episodes of low mood or anhedonia, as well as past treatment by a general practitioner or psychiatrist. As described previously, Smith et al. combined responses to these items into measures of ‘probable single episode’, ‘probable mild recurrent’ and ‘probable severe recurrent’. We identified those who strictly met all criteria for any of these categories as cases of Probable Depression. Probable Depression controls were those who met none of the criteria. Cai et al. used this definition as another minimal phenotype, but we present it here as intermediate, being more detailed than single-item measures, but less thorough than clinical assessments. A closely related second intermediate phenotype, Recurrent Depression, is included in supplementary materials.

#### Clinical phenotypes

As part of an online assessment (introduced after imaging assessments had begun), some UK Biobank participants completed the short form of the Composite International Diagnostic Interview (CIDI-SF; Kessler et al., 1998). Although not strictly a clinical assessment, the CIDI-SF is based on DSM-IV criteria. The section on MDD symptomatology provided a score of between zero and seven. Those with a score of five or more were classified as cases. However, as the assessment was completed up to 2.72 years after the imaging assessment, CIDI-assessed MDD cases who did not report any previous depressive symptoms at the time of the imaging assessment were excluded, as were controls who had previously reported symptoms. ICD-diagnosed MDD is included in supplementary materials as a secondary clinical phenotype.

### Brain imaging data

Brain magnetic resonance imaging (MRI) data were acquired at two sites, each with the same Siemens (Berlin/Munich, Germany) Skyra 3T scanner and 32-channel head coil, and using the same protocol, as described previously (Miller et al., 2016; Alfaro-Almagro et al., 2018; Smith et al., 2019). The present study used T1-weighted images, acquired using a 3D magnetisation-prepared rapid-acquisition gradient-echo (MPRAGE) sequence at 1mm isotropic resolution; T2-FLAIR images, acquired using a 3D sampling perfection with application-optimised contrasts and flip-angle evolution (SPACE) sequence at 1mm isotropic resolution and aligned to the T1 using FLIRT (Jenkinson et al., 2002); and diffusion-weighted images, acquired using a reverse phase-encoded fat-saturation sequence at 2mm isotropic resolution and modelled by the diffusion tensor.

Cortical measures were derived from raw T1 images and T2-FLAIR images (where available) by UK Biobank (Smith et al., 2019) using FreeSurfer version 6.0 (Dale et al., 1999; Fischl et al., 1999; Fischl et al., 2004). For each subject, the brain was segmented into grey matter, white matter and cerebrospinal fluid, and grey matter further segmented into cortical and subcortical regions. The cortex was then divided into regions according to the Desikan-Killany atlas (Desikan et al., 2006). Following quality control of FreeSurfer output, participants with any major errors in segmentation or cortical parcellation were excluded. For each cortical region, UK Biobank provided measures of mean thickness, surface area and volume. Prior to analysis, we summed (cortical surface area, cortical volume) or weighted-averaged (cortical thickness) metrics for some smaller regions, producing measures for 23 regions of interest per hemisphere, as detailed in supplementary materials. These were further combined to produce global and lobar (frontal, parietal, temporal, occipital and cingulate) measures.

The volumes of seven subcortical structures per hemisphere and diffusion measures – fractional anisotropy (FA) and mean diffusivity (MD) – of 27 tracts per hemisphere were previously derived using FSL by the UK Biobank imaging team (Miller et al., 2016; Alfaro-Almagro et al., 2018; Smith et al., 2019). Subcortical volumes were summed to produce a measure of overall subcortical volume. Tract-type (association fibres, thalamic radiations and projection fibres) and global summary measures of FA and MD were derived by principal components analysis, using scores on the first unrotated component (detailed in supplementary materials). Coordinates of head position within the scanner were provided by UK Biobank to be included as covariates in analyses, along with other covariates described below. Whole-brain volume (WBV) was also included as a covariate in order to reduce the effects of individual and phenotype differences in overall brain size on region-wise associations.

### Statistical analysis

Data were analysed primarily in R version 3.2.3 (R Core Team, 2013). Firstly, as an additional quality control step, outliers – defined as further than three standard deviations from the mean – were removed from all neuroimaging measures separately. We then assessed i) main effects of depression case-control status on each regional cortical metric, subcortical volume and white matter tract measure, ii) differences between depression phenotypes in each of these effects, and iii) second-level effects of phenotype on associations across all regions/tracts for each metric. Effects of case-control status were assessed using linear mixed models, including both left and right metrics with hemisphere as a random effect, and controlling age, age^2^, sex, site, scanner head position coordinates and WBV in each model. Effects of phenotype definition were then assessed using z-tests to test for significant range in β coefficients across definitions. This meant testing differences between β coefficients for the two phenotypes that showed the greatest disparity in associations with each measure, automatically accounting for any differences between phenotypes in number of included participants. Second-level analyses tested for phenotype effects on depression associations across all regions or tracts for each structural metric. FDR correction was applied to p values across all regions and depression phenotypes for each metric separately. Results are reported in terms of standardised β coefficients, with p values and q values (FDR-corrected p values) below .05 considered as significant.

## Results

Descriptive statistics and differences between depression definitions in key variables are reported in *Table 1*. As above, Ns ranged from 15,079 to 26,875, due to differences in availability of data and in criteria. Proportion of cases ranged more widely, from 9.5% for Self-reported Depression to 45.0% for Probable Depression (F_1,3_ = 118.3, p < .001) – although this was largely due to a difference between phenotypes in exclusion of controls. Mean age ranged by just one year across definitions, from 63.3 years for Probable Depression to 64.3 years for Self-reported Depression, but this was enough to produce a significant effect (F_1,3_ = 89.8, p < .001). Overall, 47.4% of subjects were male and 52.6% were female, but there was a small amount of variability in this proportion across definitions (F_1,3_ = 62.3, p < .001). There was also a small but significant effect of depression phenotype on whole-brain volume (F_1,3_ = 31.8, p = .011). Age, sex and WBV were therefore included as controls in subsequent regression models. Secondary phenotypes – Self-reported Treatment, Recurrent Depression, ICD-diagnosed MDD and Neuroticism – are summarised in supplementary materials (*Table S1*).

### Global, lobar and tract-category measures

Associations between depression phenotypes and global brain structural measures are reported in *Table 2*. Self-reported Depression was significantly associated with lower global mean cortical thickness (β = -.072, SE = .020, p < .001), lower total cortical surface area (β = -.020, SE = .008, p = .015) and lower total cortical volume (β = -.017, SE = .008, p = .041), as well as with lower global FA (β = -.084, SE = .021, p < .001) and higher global MD (β = .077, SE = .019, p < .001). Probable Depression and CIDI-assessed MDD were also both associated with lower global FA (β = -.048, SE = .017, p = .004; β = -.047, SE = .015, p = .002) and higher global MD (β = .048, SE = .016, p = .002; β = .041, SE = .014, p = .004).

**Table 2.** Associations between depression phenotypes and structural metrics for the whole brain, cortical lobes and white matter tract types.

| | Self-reported<br>Depression ($\beta$) | Probable<br>Depression ($\beta$) | CIDI-assessed<br>MDD ($\beta$) | Phenotype<br>effect (z) |
| --- | --- | --- | --- | --- |
| Global CT | **-.072\*\*\*** | -.004 | -.013 | **.069\*\*** |
| Frontal lobe CT | **-.100\*\*\*** | -.028 | **-.036\*** | **.072\*\*** |
| Parietal lobe CT | **-.052\*\*** | .007 | .011 | **.063\*\*** |
| Temporal lobe CT | **-.088\*\*\*** | -.005 | **-.034\*** | **.083\*\*** |
| Occipial lobe CT | .030 | **.045\*\*** | **.047\*\*** | .017 |
| Cingulate CT | **-.102\*\*\*** | -.019 | **-.035\*** | **.083\*\*** |
| Total CSA | **.020\*** | .007 | .004 | .016 |
| Frontal lobe CSA | **.029\*\*** | .015 | .008 | .021 |
| Parietal lobe CSA | .013 | -.001 | -.008 | .021 |
| Temporal lobe CSA | .009 | .012 | .005 | .007 |
| Occipial lobe CSA | .003 | -.008 | -.004 | .011 |
| Cingulate CSA | **.045\*\*\*** | .019 | .026** | .026 |
| Total CV | **-.017\*** | .004 | -.003 | .021 |
| Frontal lobe CV | -.021* | .001 | -.008 | .021 |
| Parietal lobe CV | -0.02 | -.001 | -.007 | .019 |
| Temporal lobe CV | -.029** | .007 | -.007 | .035** |
| Occipial lobe CV | .019 | .014 | .019 | .006 |
| Cingulate CV | .004 | .011 | .009 | .008 |
| Total SCV | -.015 | -.009 | -.012 | .007 |
| Global FA | **-.084\*\*\*** | **-.048\*\*** | **-.047\*\*** | .038 |
| Association fibres FA | **-.083\*\*\*** | **-.045\*\*** | **-.041\*\*** | .042 |
| Thalamic radiations FA | **-.092\*\*\*** | **-.047\*\*** | **-.066\*\*\*** | .045 |
| Projection fibres FA | -.014 | **-.045\*\*** | -.003 | .041 |
| Global MD | **.077\*\*\*** | **.048\*\*** | **.041\*\*** | .036 |
| Association fibres MD | **.054\*\*** | .027 | .025 | .029 |
| Thalamic radiations MD | **.092\*\*\*** | **.067\*\*\*** | **.057\*\*\*** | .035 |
| Projection fibres MD | **.061\*\*** | **.041\*** | **.041\*\*** | .019 |

*Notes.* CIDI = Composite International Diagnostic Interview (short form); MDD = Major Depressive Disorder; CT = cortical thickness; CSA = cortical surface area; CV = cortical volume;
SCV = subcortical volume; FA = fractional anisotropy; MD = mean diffusivity. Phenotype effects for each metric represent the difference, assessed using z-tests, between the two most disparate of the $\beta$ coefficients for the three phenotypes' associations with that metric. \*, \*\* and \*\*\* represent significant results at $p < .05$ , $p < .01$ and $p < .001$ , respectively; significant results after FDR correction are highlighted in bold. Corresponding structural metric associations for secondary phenotypes are reported in supplementary materials (*Table S2*).

Self-reported Depression and CIDI-assessed MDD both showed significant negative associations with most lobar cortical thickness measures (*Table 2*), while Probable Depression (β = .045, SE = .016, q = .019) and CIDI-assessed MDD (β = .047, SE = .015, q = .011) both associated with greater cortical thickness specifically in the occipital lobe. Probable Depression also showed a significant association with greater cingulate cortical surface area (β = .045, SE = .012, q = .012).

White matter tract types generally showed the same negative associations for FA and positive associations for MD with all depression phenotypes, although FA in projection fibres showed stronger associations with Probable Depression, while MD in association fibres showed stronger associations with Self-reported Depression (*Table 2*).

As the association with global mean cortical thickness was stronger for Self-reported Depression than for the other two phenotypes, there was a significant effect of phenotype definition on associations with this metric (z = .069, SE = .025, p = .007). There were also significant effects of phenotype definition on associations with cortical thickness in frontal (z = .072, SE = .025, q = .016), parietal (z = .063, SE = .024, q = .025) and temporal (z = .083, SE = .026, q = .011) lobes, as well as cingulate cortex (z = .083, SE = .026, q = .011).

As reported in supplementary materials, the three secondary depression phenotypes showed similar negative associations with cortical thickness and FA, and similar positive associations with MD and some occipital and cingulate cortex measures (*Table S2*).

### Individual regions and tracts

Depression associations for all individual cortical regions, subcortical volumes and white matter tracts are illustrated in *Figure 1*. After FDR correction, Self-reported Depression showed significant associations with lower cortical thickness in 17 of 23 regions, with the strongest associations in superior (β = -.096, SE = .018, q < .001) and inferior (β = -.093, SE = .018, q < .001) frontal gyri. Self-reported Depression also showed significant associations with higher surface area (β = .057, SE = .013, q = .002) and volume (β = .047, SE = .014, q = .034) of the cingulate isthmus, lower volume of inferior parietal (β = -.038, SE = .012, q = .034) and middle temporal (β = -.047, SE = .013, q = .023) cortices, and greater volume of the caudate nucleus (β = .060, SE = .017, q = .009) and putamen (β = .054, SE = .016, q = .010). Further, Self-reported Depression showed consistenly negative associations with white matter FA, particularly in posterior thalamic radiations (β = -.115, SE = .019, q < .001) and the forceps minor (β = -.100, SE = .020, q < .001), and consistently positive associations with MD, strongest in anterior thalmic radiations (β = .105, SE = .018, q < .001) and, again, forceps minor (β = .087, SE = .020, q < .001).

**Figure 1.**
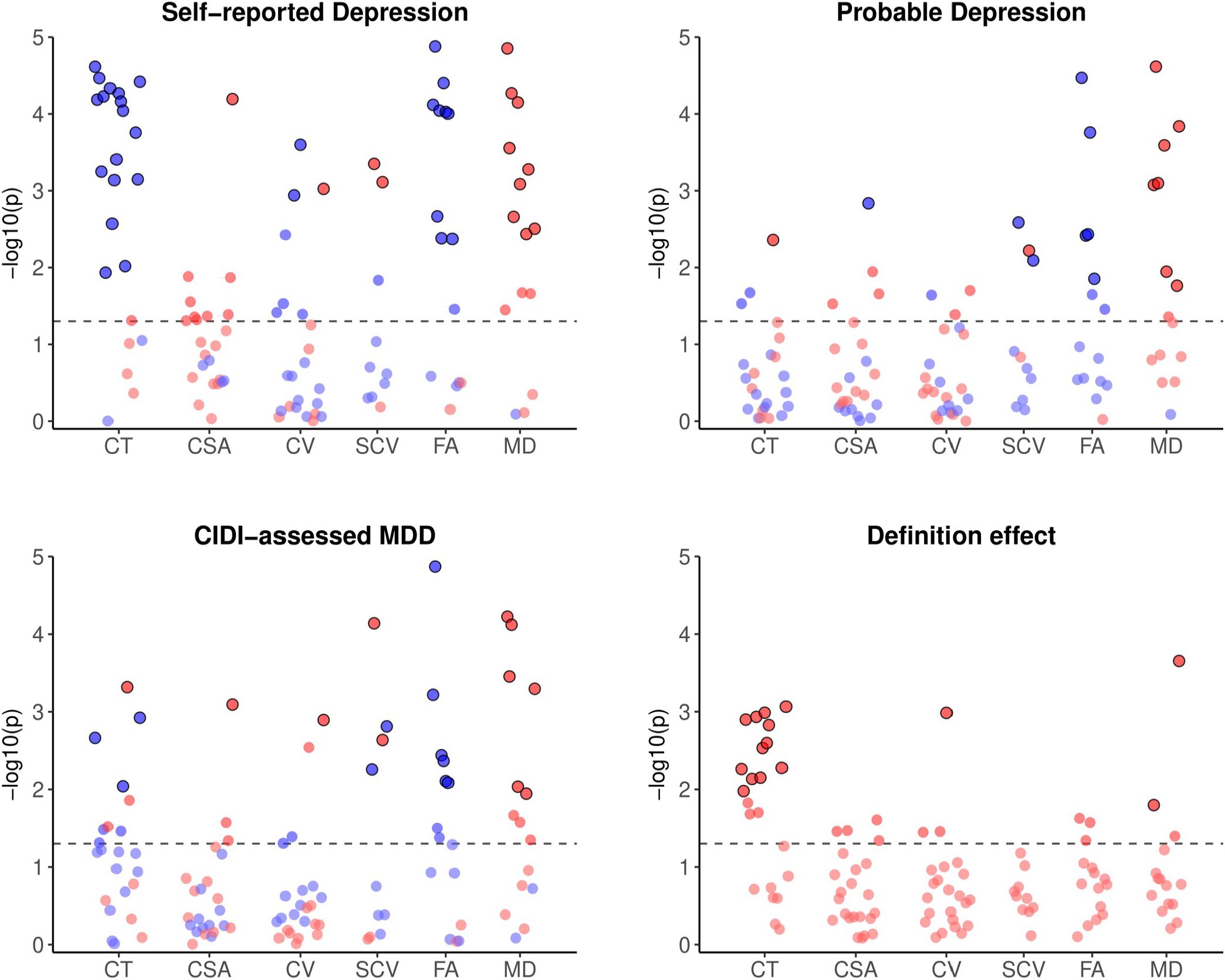
Significance of associations between depression phenotypes and individual cortical, subcortical and white matter metrics, and effects of phenotype definition on these associations. *Notes*. CIDI = Composite International Diagnostic Interview (short form); MDD = Major Depressive Disorder; CT = cortical thickness; CSA = cortical surface area; CV = cortical volume; SCV = subcortical volume; FA = fractional anisotropy; MD = mean diffusivity. Phenotype effects for each metric represent the difference, assessed using z-tests, between the two most disparate of the β coefficients for the three phenotypes’ associations with that metric. Red and blue points represent positive and negative associations, respectively. Horizontal dashed lines represent p=.05; outlined points represent associations that remained significant following FDR correction. β coefficients for all plotted results are reported in supplementary materials (*Tables S3-S8*).

Probable Depression showed weaker associations with cortical metrics, with only greater lateral occipital cortical thickness (β = .043, SE = .015, q = .025) and lower pericalcarine cortical surface area (β = -.046, SE = .014, q = .045) remaining signifiacant after correcting for multiple comparisons. Associations with lower brainstem grey matter volume (β = -.034, SE = .011, q = .018), smaller ventral diencephalon (β = -.027, SE = .010, q = .040) and larger putamen (β = .035, SE = .013, q = .034) also remained significant after FDR correction. As for Self-reported Depression, Probable Depression showed many negative associations with FA and positive associations with MD in white matter tracts, both strongest in posterior thalamic radiations (β = -.080, SE = .016, q < .001; β = .080, SE = .015, q < .001) and the forceps minor (β = -.063, SE = .017, q = .002; β = .060, SE = .017, q = .002).

Following FDR correction, CIDI-assessed MDD showed significant negative assocations with thickness of superior frontal (β = -.041, SE = .013, q = .016), fusiform (β = -.035, SE = .013, q = .041) and posterior cingulate (β = -.042, SE = .013, q = .012) cortices, and positive associations with lateral occipital cortical thickness (β = .048, SE = .014, q = .007) and cingulate isthmus surface area (β = .032, SE = .010, q = .030) and volume (β = .034, SE = .011, q = .034). CIDI-assessed MDD also showed significant associations with lower brainstem grey matter (β = -.029, SE = .011, q = .034) and ventral diencephalon (β = -.030, SE = .010, q = .014) volumes, but greater volumes of caudate nucleus (β = .055, SE = .013, q = .002) and putamen (β = .036, SE = .012, q = .018). Consistent with other phenotypes, CIDI-assessed MDD showed negative assocaitions with FA in most white matter tracts, particularly posterior (β = -.085, SE = .014, q < .001) and anterior (β = -.050, SE = .015, q = .005) thalamic radiations, and generally positive associations with MD, strongest in anterior (β = .060, SE = .013, q < .001) and superior (β = .057, SE = .013, q < .001) thalamic radiations.

Associations with lower FA and higher MD were also consistent across the secondary depression phenotypes (although not Neuroticism), while Self-reported Treatment and ICD-diagnosed MDD, as for Self-reported Depression, a number of significant associations with lower cortical thickness (*Figure S1*; *Tables S9-S14*).

### Depression definition effects

As above, associations between depression and global and lobar cortical thickness were significantly affected by definition of the depression phenotype. Significant phenotype effects were also observed at the individual region/tract level for many regional cortical thicknesses (*Figure 1*), as well as middle temporal gyrus volume (z = .054, SE = .016, q = .034), anterior thalamic radiation MD (z = .056, SE = .023, q = .046) and medial lemniscus MD (z = .072, SE = .020, q = .002). The strongest phenotype effects on association with cortical thickness were for inferior frontal (z = .076, SE = .024, q = .012), supramarginal (z = .073, SE = .023, q = .012) and fusiform (z = .073, SE = .023, q = .012) gyri (*Figure 2*). Most phenotype effects were driven by stronger structural metric associations for Self-reported Depression in particular, while the phenotype effect for medial lemniscus MD was driven by a stronger association with Probable MDD.

**Figure 2.**
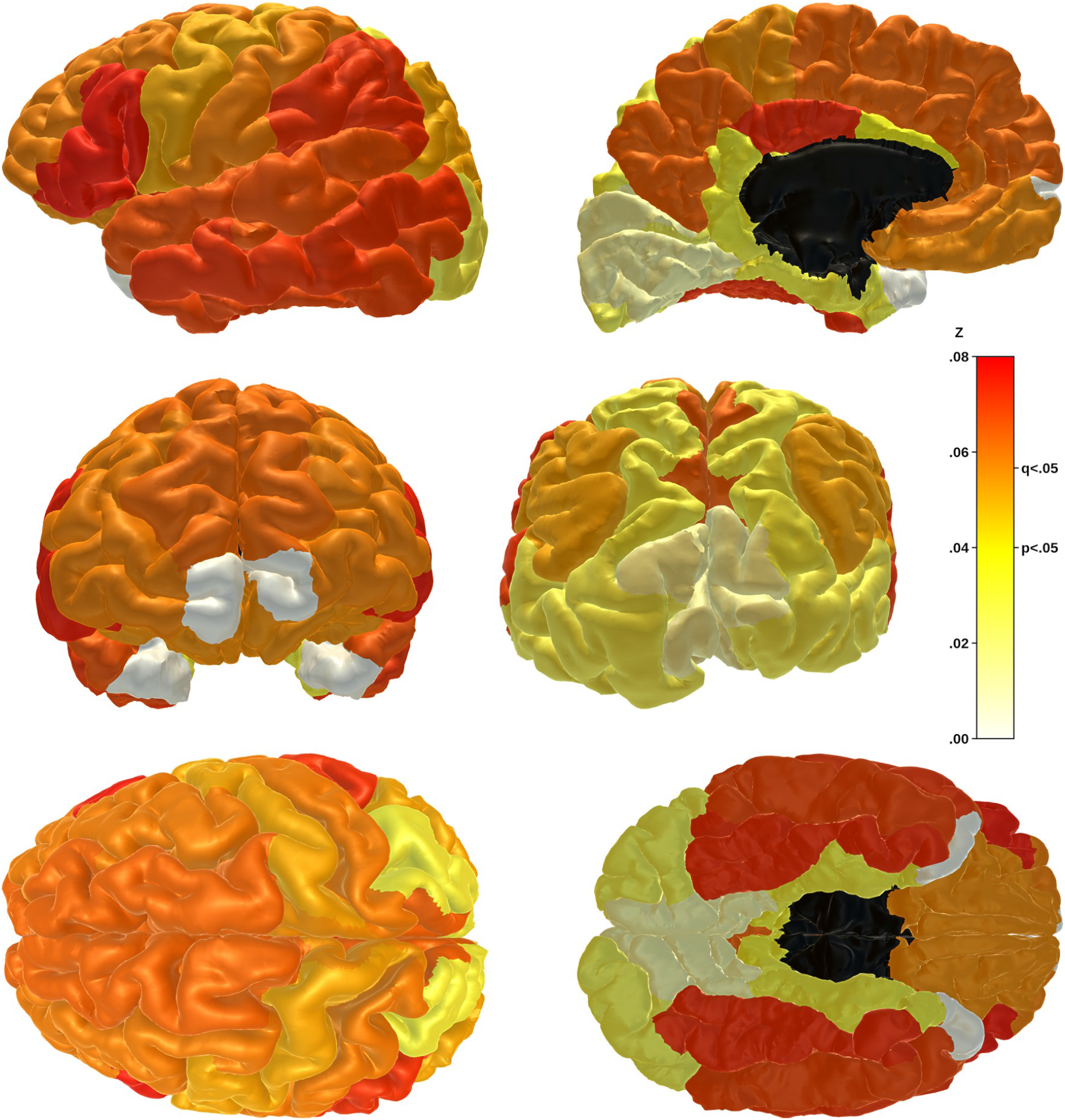
Phenotype effects on associations between depression and cortical thickness by region. *Notes*. Phenotype effects for each region represent the difference, assessed using z-tests, between the two most disparate of the β coefficients for the three phenotypes’ associations with cortical thickness of that region. Effects range from z=.00 (white) to z=.08 (red). Effects greater than z=.04 (yellow) were significant at p<.05 and those greater than z=.06 (orange) remained significant after FDR correction. Frontal and temporal pole regions were not included as regions of interest. Z statistics for plotted results are reported in supplementary materials (*Table S3*).

Second-level analyses of individual region/tract β coefficients were performed for each of the six structural metrics to assess overall effects of depression phenotype definition. These analyses confirmed a significant overall phenotype effect on associations with depression for cortical thickness (F_2,22_ = 63.47, p < .001), but also for cortical surface area (F_2,22_ = 8.73, p = .002), cortical volume (F_2,22_ = 6.15, p = .007) and white matter FA (F_2,14_ = 4.26, p = .036). Again, these effects were driven largely by stronger structural metric associations for Self-reported Depression than for the other two depression phenotypes.

## Discussion

We tested associations between three depression phenotypes and structural neuroimaging measures in the UK Biobank imaging study, and explored effects of depression phenotype definition on these associations. Cortical thickness showed more significant associations with depression than other cortical metrics, generally associating depression with thinner cortex. Self-reported Depression in particular showed robust associations with thinner cortex across most regions. Cortical surface area and volume showed few significant associations after correcting for multiple comparisons. Multiple depression phenotypes associated significantly with reduced grey matter volume in the brainstem and ventral diencephalon, but also with greater volume of caudate nucleus and putamen. For DTI-derived metrics, depression was consistently associated with lower FA and higher MD of white matter tracts. Across both measures, results for 11 of 15 tracts remained significant after correction. Results for several tracts for each measure, primarily thalamic radiations, remained significant across the three depression phenotypes.

The effects of depression definition were generally small. Across most metrics, only the volume of one cortical region and MD in two tracts showed significant phenotype definition effects after FDR correction. Cortical thickness was the exception, with 12 of 23 regions showing a significant effect of phenotype definition on depression associations. This was due to greater negative associations between cortical thickness and Self-reported Depression in particular, in contrast to the weaker and less consistently negative associations with Probable Depression and CIDI-assessed MDD. Second-level analyses confirmed a much stronger effect of phenotype definition on associations between cortical thickness and depression, but also highlighted weaker overall effects of phenotype on other metrics that were less evident at the level of indivudal regions and tracts.

Although numerous grey matter metrics did show at least nominally significant associations with depression phenotypes, these associations were generally weaker than in previous studies (Drevets et al., 2008; Koolschijn et al., 2009; Lorenzetti et al., 2009; Grieve et al., 2013). Our analyses included data for between 14,975 (Probable Depression and cortical thickness) and 26,805 (Self-reported Depression and cortical surface area) subjects, far more than most previous studies. The weaker effects may be attributable to studying lifetime MDD, rather than current MDD, or to the use of a population-based sample, rather than a clinical group. Most of the depression phenotypes were based on relatively limited self-report data, and inaccuracies in case/control classification may have diluted genuine case effects. Most associations between depression and subcortical volumes were negative, as expected, although depression phenotypes were also significantly associated with greater volumes of the caudate nucleus and putamen. These positive associations seem inconsistent with previous findings (Bora et al., 2012; Hagan et al., 2015; Lu et al., 2016), but may simply reflect lesser depression-related reduction in these subcortical structures relative to the rest of the brain, or possibly effects of antidepressant medication.

White matter measures showed more reliable results overall, with almost 65% of associations nominally significant and over 75% of those still significant after FDR correction, consistently associating depression with lower FA and higher MD. The most robust associations were for thalamic radiations, the forceps and the inferior longitudinal fasciculus. These findings are consistent with results from a previous study of a smaller subset of UK Biobank imaging participants (Shen et al., 2017), as well as studies of other cohorts (Korgaonkar et al., 2011; Ouyang et al., 2011; McIntosh et al., 2013). While cortical thickness showed more significant associations with Self-reported Depression in particular, the general contrast between grey matter and white matter metrics in terms of significant results suggests changes in connectivity might be more important in depression than changes in morphology of individual regions. This would also be consistent with studies of functional data that suggest an important role of functional dysconnectivity in depression (Wang et al., 2012; Mulders et al., 2015; Brakowski et al., 2017).

Cai et al. (2018) found that minimal phenotyping had a significant impact on depression heritability estimates and genetic associations. This is intuitively appealing and consistent with the view that less deep phenotyping, being less thorough, generates less accurate measures of depression. Conversely, our significant phenotype effects for cortical thickness were largely driven by *stronger* associations with thinner cortex for the minimal phenotype than for others. It seems therefore that minimal phenotypes may measure depression differently, rather than less accurately. Alternatively, related but opposite differences between depression phenotypes in terms of specificity and sensitivity may explain why less accurate measures produce stronger results. It is also worth noting that although Self-reported Depression was the least detailed phenotype, it was based on past *diagnosis* of depression, rather than experience of symptoms. It therefore could have been most closely associated with receiving treatment, which may at least partly account for the observed associations between depression and structural metrics. In any case, it may still be useful to study minimal phenotypes alongside more thorough assessments even when these are available.

White matter metrics, on the other hand, showed more consistent associations with depression across phenotypes and few significant effects of phenotype definition. This is more comparable to Howard et al.’s (2018) results, showing similar genetic associations for different depression definitions. Grey matter metrics other than cortical thickness also showed very few significant phenotype effects, although this may simply reflect the generally lower associations between these metrics and each depression phenotype (i.e. because associations for each phenotype were so small, differences between them were also too small to detect). Overall, our results show that whether phenotype definition impacts assocations between depression and brain structural metrics depends on which metrics are being assessed. Again, this indicates that minimal phenotypes are not simply less accurate measures, but actually differ qualitatively, which could be an important consideration for future research.

An important consideration for our results is that the depression phenotypes, by differing in terms of definition criteria, also differed in terms of the number of participants included and the proportion of cases identified. Although these differences were intrinsic to the study, they also affected the significance of associations. For example, the higher number of significant associations with cortical thickness for Self-reported Depression than for the other two phenotypes could be partly attributable to the larger number of participants with data on Self-reported Depression. However, phenotype definition effects – both for individual metrics and overall second-level analyses – considered effect sizes, rather than significance. Effect sizes would not have been affected in the same way, yet tests of phenotype effect confirmed that a greater number of signifiacant associations did correspond to stronger associations.

As already suggested, the imperfect criteria used to create most of the depression definitions was a limitation of this study. The available phenotypes showed associations with many brain structural metrics that were weak but still significant in our large sample, but more reliable measures may have produced stronger associations and more informative results. Phenotype definition effects may also have been suppressed by the relatively low associations for each depression phenotype. Another important limitation of the study might be the population-based sample. Although this ensures generalisability (despite the UK Biobank sample being slightly healthier on average than the general population of middle-aged to older adults; Fry et al., 2017), results may have been more robust if MDD cases had been recruited from a clinical population. Finally, while it was important to assess the difference between various levels of phenotyping depression as a single disorder, further research should focus on differences in brain metrics between depression subtypes, or neuroimaging associations with specific symptoms or even apparent causes of depression.

## Conclusion

We observed small but significant associations between depression at different levels of phenotyping and a range of structural neuroimaging measures. These were strongest for decreases in measures of white matter microstructural integrity, and for Self-reported Depression in particular, reduced cortical thickness. The strength of associations ranged across depression phenotypes, particularly for cortical thickness, where the minimal phenotype showed *stronger* associations than other phenotypes. This suggests that depression phenotypes may differ qualitatively in terms of the construct that they measure, rather than simply differing quantitatively in accuracy of measurement. It is therefore particularly important to consider how depression is defined when conducting and interpreting research on the disorder. Despite differences between depression phenotypes, our results also provide evidence for core neuroimaging features of depression in terms of decreased integrity of thalamic radiations and association fibres across different ways of defining the disorder.

## Data Availability

All data are available to others from UK Biobank

