## Supplementary Materials for "The Influence of Phenotyping Method on Structural Neuroimaging Associations with Depression in UK Biobank"

Mathew A. Harris[1], Simon R. Cox[2], Laura de Nooij[1], Miruna C. Barbu[1], Mark J. Adams[1], Xueyi Shen[1], Ian J. Deary[2], Stephen M. Lawrie[1], Andrew M. McIntosh[1] and Heather C. Whalley[1,*]

[1] Division of Psychiatry, University of Edinburgh, Edinburgh, UK

[2] Department of Psychology, University of Edinburgh, Edinburgh, UK

[*] corresponding author:
Heather Whalley
Division of Psychiatry, University of Edinburgh
Kennedy Tower, Royal Edinburgh Hospital
Morningside Park, Edinburgh, EH10 5HF


Running title: Neuroimaging different depression phenotypes

Keywords: Depression; structural neuroimaging; levels of phenotyping; grey matter; white matter integrity; UK Biobank

Supplementary word count: 508
Supplementary figures: 1
Supplementary tables: 14

### Methods

#### Secondary phenotypes

*Self-reported Treatment:* Also derived from a single self-report item, but based on whether participants reported that they had (cases) or had not (controls) ever sought treatment from a general practitioner or psychiatrist for symptoms of ‘nerves, anxiety or depression’. This was not included in our main analyses as it represents a very loose definition of depression.

*Recurrent Depression:* Closely related to Probable Depression, based on the same criteria, but with subjects previously classified as ‘probable single episode’ excluded. This was not included in our main analyses due to similarity to Probable Depression. This phenotype was also not included in Cai et al.’s (2018) study.

*ICD-diagnosed MDD:* A more stringent clinical definition, derived from hospital records of an ICD-9/10 diagnosis provided by a clinician. This phenotype was not included in main analyses as it did not cover a full lifetime history of depression, only records of depression at times when participants had been in hospital. It therefore identified relatively few cases compared to other phenotypes.

*Neuroticism:* A personality trait closely related to depression, Neuroticims was derived from the 12-item neuroticism scale of the Eysenck Personality Questionnaire Revised (EPQ-R; Eysenck, Eysenck & Barrett, 1985), administered by touchscreen at UK Biobank assessment centres. Scores ranged from 0 to 12, with scores of >8 considered as representing high Neuroticism.

Descriptive statistics for secondary depression phenotypes are presented in *Table S1*, results are summarised in *Table S2* and *Figure S1*, and full results by tract/region are reported in *Tables S9-S14*.

#### Brain imaging data

The following cortical regions of interest were combinations of Desikan-Killany atlas regions: middle frontal gyrus (rostral middle frontal gyrus, caudal middle frontal gyrus), inferior frontal gyrus (pars orbitalis, pars triangularis, pars opercularis), orbitofrontal cortex (lateral orbitofrontal cortex, medial orbitofrontal cortex), superior temporal gyrus (superior temporal gyrus, transverse temporal gyrus, banks of superior temporal sulcus), and medial temporal cortex (parahippocampal cortex, entorhinal cortex). Additionally, the parcellated region paracentral cortex was divided in two and half added to the precentral and postcentral gyri. Parcellated regions frontal pole and temporal pole were not included as regions of interest, but were included in global and lobar measures.

Lobar measures were combinations of metrics from both left and right regions, as follows: frontal (superior frontal gyrus, middle frontal gyrus, inferior frontal gyrus, orbitofrontal cortex, precentral gyrus, frontal pole), parietal (postcentral gyrus, superior parietal cortex, inferior parietal cortex, supramarginal gyrus, precuneus), temporal (insula, superior temporal gyrus, middle temporal gyrus, inferior temporal gyrus, fusiform gyrus, medial temporal cortex, temporal pole), occipital (lateral occipital cortex, cuneus, pericalcarine cortex, lingual gyrus) and cingulate (rostral anterior cingulate cortex, caudal anterior cingulate cortex, posterior cingulate cortex, cingulate isthmus).

White matter microstructural metrics were combined by principal component analysis, taking scores on the first unrotated principal component. For global FA, this component explained 41.2% of variance in individual tract FA measures, with tract-type components explaining 44.9% (association fibres), 58.0% (thalamic radiations) and 49.5% (projection fibres) of variance. Components used as summary measures of MD explained 41.9% (global), 47.6% (association fibres), 62.1% (thalamic radiations) and 54.1% (projection fibres) of variance.

**Table S1 – Descriptive statistics for secondary phenotypes**

| | Self-reported Treatment | Recurrent Depression | ICD-diagnosed MDD | High Neuroticism |
|---|---|---|---|---|
| Subjects (N, %) | 38,791 (98.7) | 13,473 (34.3) | 17,167 (43.7) | 33,190 (84.5) |
| Age (years; M, SD) | 64.1 (7.5) | 63.4 (7.6) | 64.9 (7.7) | 64.1 (7.5) |
| Sex | | | | |
| Males (N, %) | 18,404 (47.4) | 6,565 (48.7) | 8,919 (52.0) | 15,924 (48.0) |
| Females (N, %) | 20,387 (52.6) | 6,908 (51.3) | 8,248 (48.0) | 17,266 (52.0) |
| Depression | | | | |
| Cases (N, %) | 14,521 (37.4) | 5,181 (38.5) | 554 (3.2) | 27,345 (82.4) |
| Controls (N, %) | 24,270 (62.6) | 8,292 (61.5) | 16,613 (96.8) | 5,845 (17.6) |
| Neuroticism score (M, SD) | 3.8 (3.2) | 3.8 (3.2) | 3.1 (2.8) | 3.8 (3.2) |
| Global CT (mm; M, SD) | 2.67 (.11) | 2.67 (.11) | 2.67 (.11) | 2.67 (.11) |
| Global CSA ($mm^2$; M, SD) | 169,074 (15,384) | 169,593 (15,295) | 169,485 (15,311) | 169,304 (15,400) |
| Global CV ($mm^3$; M, SD) | 498,214 (47,723) | 499,694 (47,539) | 498,654 (47,417) | 498,893 (47,754) |
| Total SCV ($mm^3$; M, SD) | 187,727 (16,955) | 188,154 (16,856) | 187,958 (16,819) | 187,945 (16,972) |
| Global FA (M, SD) | .000 (1.000) | .029 (.992) | -.017 (1.010) | .006 (1.001) |
| Global MD (M, SD) | -.001 (1.000) | .005 (.990) | .051 (1.018) | -.006 (1.000) |

*Notes*. ICD = International Classification of Diseases (9/10); MDD = Major Depressive Disorder; CT = cortical thickness; CSA = cortical surface area; CV = cortical volume; SCV = subcortical volume; FA = fractional anisotropy; MD = mean diffusivity. Descriptive statistics for primary depression phenotypes are reported in *Table 1*.

**Table S2 – Associations between secondary phenotypes and structural metrics for the whole brain, cortical lobes and white matter tract types**

| | Self-reported Treatment (β) | Recurrent Depression (β) | ICD-diagnosed MDD (β) | Neuroticism score (β) |
|---|---|---|---|---|
| Global CT | -.018 | -.001 | -.058 | .012 |
| Frontal lobe CT | **-.031**** | -.030 | **-.099*** | -.001 |
| Parietal lobe CT | -.007 | .011 | -.018 | .027 |
| Temporal lobe CT | **-.026**** | -.004 | **-.118**** | .008 |
| Occipial lobe CT | **.026*** | **.053**** | .084* | **.044*** |
| Cingulate CT | **-.032**** | -.023 | **-.104*** | -.004 |
| Total CSA | .007 | .006 | .024 | **-.014*** |
| Frontal lobe CSA | .012* | .014 | .046* | -.015 |
| Parietal lobe CSA | .004 | -.001 | -.016 | -.014 |
| Temporal lobe CSA | .003 | .012 | .028 | -.002 |
| Occipial lobe CSA | -.001 | -.016 | .012 | -.027 |
| Cingulate CSA | **.020**** | .022* | .054* | -.012 |
| Total CV | -.001 | .004 | -.007 | .001 |
| Frontal lobe CV | -.003 | -.001 | -.006 | -.006 |
| Parietal lobe CV | -.003 | .001 | -.025 | .002 |
| Temporal lobe CV | -.007 | .009 | -.027 | .009 |
| Occipial lobe CV | .012 | .012 | .057 | -.003 |
| Cingulate CV | .006 | .012 | -.003 | -.016 |
| Total SCV | -.006 | -.010 | -.047 | -.013 |
| Global FA | **-.026*** | **-.061***** | **-.112**** | -.004 |
| Association fibres FA | **-.024*** | **-.057**** | **-.110**** | .001 |
| Thalamic radiations FA | **-.030**** | **-.064***** | **-.101*** | -.023 |
| Projection fibres FA | -.017 | **-.038*** | **-.089*** | -.009 |
| Global MD | **.022*** | **.054**** | **.140***** | .017 |
| Association fibres MD | .005 | .031 | **.122**** | .006 |
| Thalamic radiations MD | **.033***** | **.075***** | **.148***** | .023 |
| Projection fibres MD | **.033**** | **.042*** | **.118**** | .026 |

*Notes*. CIDI = Composite International Diagnostic Interview (short form); MDD = Major Depressive Disorder; CT = cortical thickness; CSA = cortical surface area; CV = cortical volume; SCV = subcortical volume; FA = fractional anisotropy; MD = mean diffusivity. *, ** and *** represent significant results at $p<.05$, $p<.01$ and $p<.001$, respectively; significant results after FDR correction are highlighted in bold. Corresponding structural metric associations for primary depression phenotypes and phenotype effects are reported in *Table 2*.

**Figure S1 – Significance of associations between secondary phenotypes and individual cortical, subcortical and white matter metrics**

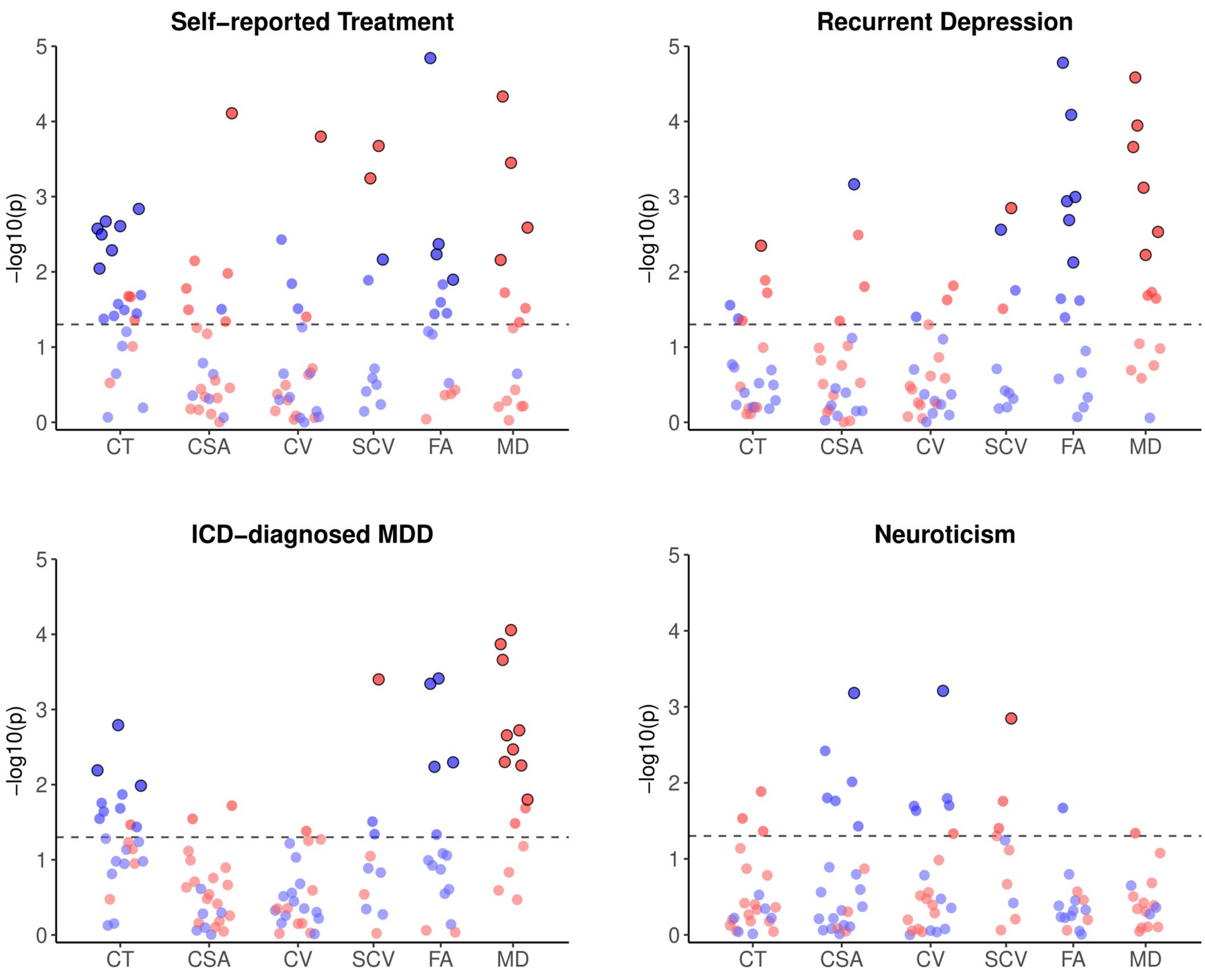


*Notes*. ICD = International Classification of Diseases (9/10); MDD = Major Depressive Disorder; CT = cortical thickness; CSA = cortical surface area; CV = cortical volume; SCV = subcortical volume; FA = fractional anisotropy; MD = mean diffusivity. Red and blue points represent positive and negative associations, respectively. Horizontal dashed lines represent p=.05; outlined points represent associations that remained significant following FDR correction. β coefficients for all plotted results are reported in *Tables S9-S14*.

**Table S3 – Associations between depression phenotypes and cortical thickness by region**

| | Self-reported Depression (β) | Probable Depression (β) | CIDI-assessed MDD (β) | Phenotype effect (z) |
|---|---|---|---|---|
| Superior frontal gyrus | **-.096*** | -.032* | **-.041** | **.064** |
| Middle frontal gyrus | **-.080*** | -.020 | -.025 | **.060*** |
| Inferior frontal gyrus | **-.093*** | -.016 | -.027* | **.076** |
| Orbitofrontal cortex | **-.063*** | -.006 | -.026 | .058* |
| Precentral gyrus | **-.082*** | -.034* | -.029* | .053* |
| Postcentral gyrus | **-.045*** | .013 | .015 | **.060** |
| Superior parietal cortex | .000 | .018 | .030* | .030 |
| Supramarginal gyrus | **-.086*** | -.011 | -.012 | **.073** |
| Inferior parietal cortex | **-.054** | -.002 | -.002 | .053* |
| Precuneus | **-.063*** | .002 | -.001 | **.065** |
| Superior temporal gyrus | **-.063*** | .005 | -.021 | **.068** |
| Middle temporal gyrus | **-.078*** | -.006 | -.024 | **.072** |
| Inferior temporal gyrus | **-.077*** | -.008 | -.028* | **.069** |
| Fusiform gyrus | **-.072*** | .001 | **-.035** | **.073** |
| Medial temporal cortex | **-.047** | -.022 | -.017 | .030 |
| Lateral occipital cortex | .022 | **.043** | **.048*** | .027 |
| Cuneus | .030 | .021 | .033* | .012 |
| Pericalcarine cortex | .036* | .028 | .010 | .026 |
| Lingual cortex | .014 | .025 | .019 | .011 |
| Rostral ACC | **-.063*** | -.003 | -.023 | **.060** |
| Caudal ACC | **-.053*** | -.014 | -.018 | .039 |
| Posterior cingulate cortex | **-.085*** | -.011 | **-.042** | **.074*** |
| Cingulate isthmus | -.030 | -.007 | .003 | .033 |

*Notes*. CIDI = Composite International Diagnostic Interview (short form); MDD = Major Depressive Disorder; ACC = anterior cingulate cortex. *, ** and *** represent significant results at p<.05, p<.01 and p<.001, respectively; significant results after FDR correction are highlighted in bold.

**Table S4 – Associations between depression phenotypes and cortical surface area by region**

| | Self-reported Depression (β) | Probable Depression (β) | CIDI-assessed MDD (β) | Phenotype effect (z) |
|---|---|---|---|---|
| Superior frontal gyrus | .020* | .018* | .011 | .009 |
| Middle frontal gyrus | .032* | .016 | .007 | .024 |
| Inferior frontal gyrus | .031* | .010 | -.006 | .037* |
| Orbitofrontal cortex | .013 | -.004 | .000 | .017 |
| Precentral gyrus | .025* | .005 | .012 | .020 |
| Postcentral gyrus | .024* | .006 | -.004 | .027 |
| Superior parietal cortex | .008 | -.004 | -.008 | .016 |
| Supramarginal gyrus | .022 | .006 | -.013 | .035* |
| Inferior parietal cortex | -.016 | -.011 | -.005 | .011 |
| Precuneus | .019 | -.004 | .003 | .023 |
| Superior temporal gyrus | .023* | .018 | .012 | .011 |
| Middle temporal gyrus | -.018 | .009 | -.006 | .027 |
| Inferior temporal gyrus | .001 | -.002 | -.003 | .004 |
| Fusiform gyrus | .013 | .000 | .004 | .013 |
| Medial temporal cortex | .023 | .019 | .020 | .004 |
| Lateral occipital cortex | .014 | .009 | .012 | .005 |
| Cuneus | .017 | -.018 | -.011 | .034 |
| Pericalcarine cortex | -.018 | **-.046**** | -.024 | .028 |
| Lingual cortex | -.017 | -.002 | -.007 | .016 |
| Rostral ACC | .016 | .018* | .015* | .003 |
| Caudal ACC | .020* | .009 | .015* | .011 |
| Posterior cingulate cortex | .033* | -.005 | .005 | .038* |
| Cingulate isthmus | **.057***** | .024* | **.032***** | .033* |

*Notes*. CIDI = Composite International Diagnostic Interview (short form); MDD = Major Depressive Disorder; ACC = anterior cingulate cortex. *, ** and *** represent significant results at p<.05, p<.01 and p<.001, respectively; significant results after FDR correction are highlighted in bold.

**Table S5 – Associations between depression phenotypes and cortical volume by region**

| | Self-reported Depression (β) | Probable Depression (β) | CIDI-assessed MDD (β) | Phenotype effect (z) |
|---|---|---|---|---|
| Superior frontal gyrus | -.022* | .007 | -.005 | .029* |
| Middle frontal gyrus | .002 | .011 | .002 | .009 |
| Inferior frontal gyrus | -.005 | .010 | -.008 | .018 |
| Orbitofrontal cortex | -.026* | -.013 | -.018* | .013 |
| Precentral gyrus | **-.039**** | -.025* | -.012 | .027 |
| Postcentral gyrus | -.014 | .008 | .004 | .023 |
| Superior parietal cortex | .007 | .002 | .004 | .005 |
| Supramarginal gyrus | -.015 | .001 | -.020* | .021 |
| Inferior parietal cortex | **-.038**** | -.010 | -.007 | .031* |
| Precuneus | -.005 | -.003 | .000 | .006 |
| Superior temporal gyrus | -.008 | .018 | .002 | .026 |
| Middle temporal gyrus | **-.047***** | .007 | -.010 | **.054**** |
| Inferior temporal gyrus | -.027* | -.005 | -.013 | .021 |
| Fusiform gyrus | -.018 | -.003 | -.007 | .015 |
| Medial temporal cortex | -.003 | .003 | .012 | .015 |
| Lateral occipital cortex | .022 | .023* | .031** | .009 |
| Cuneus | .030 | -.004 | .012 | .035 |
| Pericalcarine cortex | .000 | -.027 | -.018 | .027 |
| Lingual cortex | .004 | .012 | .008 | .008 |
| Rostral ACC | -.005 | .014 | .002 | .019 |
| Caudal ACC | -.010 | .000 | .005 | .015 |
| Posterior cingulate cortex | -.002 | -.007 | -.012 | .010 |
| Cingulate isthmus | **.047***** | .027* | **.034**** | .020 |

*Notes*. CIDI = Composite International Diagnostic Interview (short form); MDD = Major Depressive Disorder; ACC = anterior cingulate cortex. *, ** and *** represent significant results at $p<.05$, $p<.01$ and $p<.001$, respectively; significant results after FDR correction are highlighted in bold.

**Table S6 – Associations between depression phenotypes and subcortical volumes**

| | Self-reported Depression (β) | Probable Depression (β) | CIDI-assessed MDD (β) | Phenotype effect (z) |
|---|---|---|---|---|
| Amygdala | -.017 | -.005 | .003 | .019 |
| Brainstem | -.010 | **-.034**** | **-.029**** | .024 |
| Caudate nucleus | **.060***** | .020 | **.055***** | .035 |
| Cerebellum | -.025 | -.008 | -.015 | .018 |
| Hippocampus | -.036* | -.004 | -.009 | .031 |
| Nucleus accumbens | -.009 | -.018 | .002 | .020 |
| Pallidum | .007 | -.015 | -.004 | .022 |
| Putamen | **.054***** | **.035**** | **.036**** | .018 |
| Thalamus | -.012 | -.011 | -.008 | .005 |
| Ventral diencephalon | -.015 | **-.027**** | **-.030**** | .015 |

*Notes*. CIDI = Composite International Diagnostic Interview (short form); MDD = Major Depressive Disorder. *, ** and *** represent significant results at $p<.05$, $p<.01$ and $p<.001$, respectively; significant results after FDR correction are highlighted in bold.

**Table S7 – Associations between depression phenotypes and fractional anisotropy by tract**

| | Self-reported Depression (β) | Probable Depression (β) | CIDI-assessed MDD (β) | Phenotype effect (z) |
|---|---|---|---|---|
| Acoustic radiation | -.020 | -.015 | -.021 | .005 |
| Anterior thalamic radiation | **-.083*** ** | -.026 | **-.050*** ** | .057* |
| Posterior thalamic radiation | **-.115*** ** | **-.080*** ** | **-.085*** ** | .035 |
| Superior thalamic radiation | **-.063** ** | -.018 | -.033* | .045 |
| Cingulum cingulate | **-.065*** ** | **-.038** ** | -.024* | .040* |
| Forceps major | **-.062** ** | **-.051** ** | **-.047** ** | .015 |
| Forceps minor | **-.100*** ** | **-.063*** ** | **-.044** ** | .057* |
| Inferior FO fasciculus | **-.076*** ** | -.036* | **-.038** ** | .041 |
| Inferior longitudinal fasciculus | **-.076*** ** | **-.039* ** | **-.038** ** | .038 |
| Parahippocampal cingulate | .007 | -.010 | -.002 | .017 |
| Superior longitudinal fasciculus | **-.053** ** | -.021 | -.026 | .031 |
| Uncinate fasciculus | -.039* | -.015 | -.021 | .024 |
| Corticospinal tract | -.018 | .001 | .002 | .020 |
| Medial lemniscus | -.018 | -.030* | -.002 | .029 |
| Middle cerebellar peduncle | .021 | -.016 | .009 | .038 |

*Notes*. CIDI = Composite International Diagnostic Interview (short form); MDD = Major Depressive Disorder; FO = fronto-occipital. *, ** and *** represent significant results at p<.05, p<.01 and p<.001, respectively; significant results after FDR correction are highlighted in bold.

**Table S8 – Associations between depression phenotypes and mean diffusivity by tract**

| | Self-reported Depression (β) | Probable Depression (β) | CIDI-assessed MDD (β) | Phenotype effect (z) |
|---|---|---|---|---|
| Acoustic radiation | .038* | .021 | .011 | .027 |
| Anterior thalamic radiation | **.105*** ** | **.049*** ** | **.060*** ** | **.056*** |
| Posterior thalamic radiation | **.066*** ** | **.080*** ** | **.049*** ** | .031 |
| Superior thalamic radiation | **.083*** ** | **.049*** ** | **.057*** ** | .034 |
| Cingulum cingulate | **.059** ** | .023 | .033* | .036 |
| Forceps major | -.005 | .017 | -.004 | .021 |
| Forceps minor | **.087*** ** | **.060*** ** | **.039** ** | .047 |
| Inferior FO fasciculus | **.063*** ** | **.039*** | .031* | .032 |
| Inferior longitudinal fasciculus | .044* | .031* | .019 | .024 |
| Parahippocampal cingulate | .005 | -.003 | .007 | .010 |
| Superior longitudinal fasciculus | **.057** ** | .031 | **.037*** | .026 |
| Uncinate fasciculus | **.064*** ** | .015 | .022 | .049* |
| Corticospinal tract | .045* | **.038*** | .029* | .016 |
| Medial lemniscus | .013 | **.055*** ** | -.017 | **.072*** ** |
| Middle cerebellar peduncle | **.062** ** | .025 | **.054*** ** | .037 |

*Notes*. CIDI = Composite International Diagnostic Interview (short form); MDD = Major Depressive Disorder; FO = fronto-occipital. *, ** and *** represent significant results at p<.05, p<.01 and p<.001, respectively; significant results after FDR correction are highlighted in bold.

**Table S9 – Associations between secondary phenotypes and cortical thickness by region**

| | Self-reported Treatment (β) | Recurrent Depression (β) | ICD-diagnosed MDD (β) | Neuroticism score (β) |
|---|---|---|---|---|
| Superior frontal gyrus | **-.027**** | -.035* | -.100** | .005 |
| Middle frontal gyrus | **-.024**** | -.022 | -.082* | .008 |
| Inferior frontal gyrus | **-.028**** | -.021 | -.090* | -.008 |
| Orbitofrontal cortex | -.019* | -.009 | -.086* | .002 |
| Precentral gyrus | **-.029**** | -.033* | -.073 | -.002 |
| Postcentral gyrus | -.002 | .015 | -.012 | .028 |
| Superior parietal cortex | .010 | .033* | .036 | .035* |
| Supramarginal gyrus | **-.026**** | -.013 | -.053 | .014 |
| Inferior parietal cortex | -.019* | .004 | -.014 | .024 |
| Precuneus | -.011 | .007 | -.062 | .010 |
| Superior temporal gyrus | -.020* | .005 | **-.114**** | .007 |
| Middle temporal gyrus | **-.026**** | -.007 | -.081* | -.001 |
| Inferior temporal gyrus | -.015 | -.008 | -.091* | .013 |
| Fusiform gyrus | -.020* | .008 | -.059 | .012 |
| Medial temporal cortex | -.017 | -.016 | -.066 | -.016 |
| Lateral occipital cortex | .022* | **.046**** | .072 | .040* |
| Cuneus | .021* | .026 | .078* | .032* |
| Pericalcarine cortex | .015 | .039* | .067 | -.012 |
| Lingual cortex | .019* | .037* | .059 | .022 |
| Rostral ACC | -.018* | -.006 | -.072* | .006 |
| Caudal ACC | **-.025**** | -.017 | -.061 | -.007 |
| Posterior cingulate cortex | -.020* | -.015 | -.091* | .002 |
| Cingulate isthmus | -.004 | -.010 | -.058 | .012 |

*Notes*. ICD = International Classification of Diseases (9/10); MDD = Major Depressive Disorder; ACC = anterior cingulate cortex. *, ** and *** represent significant results at p<.05, p<.01 and p<.001, respectively; significant results after FDR correction are highlighted in bold.

**Table S10 – Associations between secondary phenotypes and cortical surface area by region**

| | Self-reported Treatment (β) | Recurrent Depression (β) | ICD-diagnosed MDD (β) | Neuroticism score (β) |
|---|---|---|---|---|
| Superior frontal gyrus | .012* | .014 | .025 | -.004 |
| Middle frontal gyrus | .014* | .016 | .047 | -.012 |
| Inferior frontal gyrus | .003 | .012 | .047 | -.002 |
| Orbitofrontal cortex | -.004 | -.001 | .052* | -.029** |
| Precentral gyrus | .017** | .004 | .034 | -.027* |
| Postcentral gyrus | .012 | .005 | -.004 | -.016 |
| Superior parietal cortex | .003 | -.007 | .012 | -.003 |
| Supramarginal gyrus | .006 | .009 | -.032 | -.006 |
| Inferior parietal cortex | -.008 | -.010 | -.016 | -.025* |
| Precuneus | .005 | -.003 | -.007 | .003 |
| Superior temporal gyrus | .010 | .020* | .023 | .000 |
| Middle temporal gyrus | -.005 | .015 | .028 | -.008 |
| Inferior temporal gyrus | .002 | .000 | .000 | -.004 |
| Fusiform gyrus | -.008 | -.009 | .036 | .001 |
| Medial temporal cortex | .008 | .021 | .008 | .008 |
| Lateral occipital cortex | .005 | .001 | .026 | -.004 |
| Cuneus | .000 | -.024 | .014 | -.036** |
| Pericalcarine cortex | -.019* | **-.053*** ** | -.024 | **-.053*** ** |
| Lingual cortex | -.001 | -.005 | .004 | -.020 |
| Rostral ACC | .009* | .023** | .028 | -.016* |
| Caudal ACC | .013* | .009 | .025 | -.010 |
| Posterior cingulate cortex | .006 | -.004 | .016 | -.009 |
| Cingulate isthmus | **.027*** ** | .027* | .063* | .017 |

*Notes*. ICD = International Classification of Diseases (9/10); MDD = Major Depressive Disorder; ACC = anterior cingulate cortex. *, ** and *** represent significant results at p<.05, p<.01 and p<.001, respectively; significant results after FDR correction are highlighted in bold.

**Table S11 – Associations between secondary phenotypes and cortical volume by region**

| | Self-reported Treatment (β) | Recurrent Depression (β) | ICD-diagnosed MDD (β) | Neuroticism score (β) |
|---|---|---|---|---|
| Superior frontal gyrus | .002 | .002 | -.016 | .004 |
| Middle frontal gyrus | .005 | .011 | .019 | .000 |
| Inferior frontal gyrus | -.005 | .011 | .002 | .002 |
| Orbitofrontal cortex | -.018** | -.014 | -.010 | -.025* |
| Precentral gyrus | -.008 | -.024* | -.028 | -.027* |
| Postcentral gyrus | .006 | .007 | -.016 | .002 |
| Superior parietal cortex | .005 | .007 | .023 | .013 |
| Supramarginal gyrus | -.005 | .002 | -.052 | .001 |
| Inferior parietal cortex | -.014* | -.008 | -.026 | -.014 |
| Precuneus | .001 | .000 | -.024 | .011 |
| Superior temporal gyrus | .001 | .021 | -.043 | .012 |
| Middle temporal gyrus | -.014* | .013 | .010 | -.002 |
| Inferior temporal gyrus | -.001 | -.003 | -.034 | .009 |
| Fusiform gyrus | -.013 | -.007 | .011 | .008 |
| Medial temporal cortex | .000 | .008 | -.026 | -.001 |
| Lateral occipital cortex | .015* | .018 | .058* | .020 |
| Cuneus | .010 | -.008 | .062 | -.013 |
| Pericalcarine cortex | -.011 | -.027 | .003 | **-.053*** ** |
| Lingual cortex | .011 | .017 | .039 | -.003 |
| Rostral ACC | .001 | .019* | -.001 | -.021* |
| Caudal ACC | -.002 | -.003 | -.016 | -.023* |
| Posterior cingulate cortex | -.001 | -.010 | -.015 | -.010 |
| Cingulate isthmus | **.027*** ** | .030* | .057 | .025* |

*Notes*. ICD = International Classification of Diseases (9/10); MDD = Major Depressive Disorder; ACC = anterior cingulate cortex. *, ** and *** represent significant results at p<.05, p<.01 and p<.001, respectively; significant results after FDR correction are highlighted in bold.

**Table S12 – Associations between secondary phenotypes and subcortical volumes**

| | Self-reported Treatment (β) | Recurrent Depression (β) | ICD-diagnosed MDD (β) | Neuroticism score (β) |
|---|---|---|---|---|
| Amygdala | -.006 | -.005 | -.020 | .023* |
| Brainstem | -.018* | **-.037**** | -.045 | .002 |
| Caudate nucleus | **.030***** | .032* | .060 | .036* |
| Cerebellum | -.009 | -.012 | -.067* | -.025 |
| Hippocampus | -.010 | -.006 | -.060* | .016 |
| Nucleus accumbens | -.003 | -.016 | .031 | .024 |
| Pallidum | -.007 | -.011 | .002 | .023 |
| Putamen | **.030***** | **.045**** | **.117***** | **.044**** |
| Thalamus | -.004 | -.008 | -.038 | -.010 |
| Ventral diencephalon | **-.017**** | -.027* | -.017 | .006 |

*Notes*. ICD = International Classification of Diseases (9/10); MDD = Major Depressive Disorder. *, ** and *** represent significant results at p<.05, p<.01 and p<.001, respectively; significant results after FDR correction are highlighted in bold.

**Table S13 – Associations between secondary phenotypes and fractional anisotropy by tract**

| | Self-reported Treatment (β) | Recurrent Depression (β) | ICD-diagnosed MDD (β) | Neuroticism score (β) |
|---|---|---|---|---|
| Acoustic radiation | .001 | -.018 | .006 | -.013 |
| Anterior thalamic radiation | -.018 | -.039* | -.067 | -.009 |
| Posterior thalamic radiation | **-.057*** ** | **-.097*** ** | **-.141*** ** | -.039* |
| Superior thalamic radiation | -.019 | -.037* | -.066 | -.009 |
| Cingulum cingulate | -.017* | **-.046** ** | **-.092** ** | .002 |
| Forceps major | **-.030** ** | **-.058** ** | -.090* | -.027 |
| Forceps minor | **-.030** ** | **-.075*** ** | **-.150*** ** | -.010 |
| Inferior FO fasciculus | -.022* | **-.045** ** | -.060 | -.012 |
| Inferior longitudinal fasciculus | -.024* | **-.056** ** | -.070 | -.016 |
| Parahippocampal cingulate | .007 | -.003 | -.041 | .018 |
| Superior longitudinal fasciculus | -.020* | -.036* | -.065 | -.002 |
| Uncinate fasciculus | -.010 | -.020 | -.044 | .000 |
| Corticospinal tract | .008 | -.008 | -.014 | .016 |
| Medial lemniscus | **-.023*** | -.025 | **-.104** ** | -.011 |
| Middle cerebellar peduncle | .010 | -.014 | .004 | .009 |

*Notes*. ICD = International Classification of Diseases (9/10); MDD = Major Depressive Disorder; FO = fronto-occipital. *, ** and *** represent significant results at p<.05, p<.01 and p<.001, respectively; significant results after FDR correction are highlighted in bold.

**Table S14 – Associations between secondary phenotypes and mean diffusivity by tract**

| | Self-reported Treatment (β) | Recurrent Depression (β) | ICD-diagnosed MDD (β) | Neuroticism score (β) |
|---|---|---|---|---|
| Acoustic radiation | .005 | .020 | .043 | -.019 |
| Anterior thalamic radiation | **.025**** | **.059***** | **.142***** | .016 |
| Posterior thalamic radiation | **.044***** | **.085***** | **.138***** | .032* |
| Superior thalamic radiation | .022* | **.061***** | **.105**** | .012 |
| Cingulum cingulate | .006 | .028 | **.121**** | .002 |
| Forceps major | .001 | .021 | .064 | .005 |
| Forceps minor | **.037***** | **.060***** | **.169***** | .016 |
| Inferior FO fasciculus | .018 | **.046**** | **.114**** | .011 |
| Inferior longitudinal fasciculus | .009 | .039* | .083* | .005 |
| Parahippocampal cingulate | -.011 | -.003 | .036 | -.010 |
| Superior longitudinal fasciculus | .020* | .041* | **.125**** | .022 |
| Uncinate fasciculus | .005 | .022 | **.105**** | .013 |
| Corticospinal tract | .005 | .040* | .075 | -.013 |
| Medial lemniscus | .019* | **.046**** | .084* | .004 |
| Middle cerebellar peduncle | **.032**** | .030 | **.104*** | .032 |

*Notes*. ICD = International Classification of Diseases (9/10); MDD = Major Depressive Disorder; FO = fronto-occipital. *, ** and *** represent significant results at $p<.05$, $p<.01$ and $p<.001$, respectively; significant results after FDR correction are highlighted in bold.
